# Precise interpretation and prioritization of sequence variants with Onkopus for supporting molecular tumor boards

**DOI:** 10.1101/2024.09.24.24314298

**Authors:** Nadine S. Kurz, Kevin Kornrumpf, Tim Tucholski, Klara Drofenik, Alexander König, Tim Beißbarth, Jürgen Dönitz

## Abstract

One of the major challenges in precision medicine is the identification of pathogenic, actionable variants and the selection of personalized treatments. We present Onkopus, a variant interpretation framework, based on a modular architecture, for interpreting and prioritizing genetic alterations in cancer patients. We show that aggregation and harmonization of clinical databases, coupled with querying of these databases to varying associated biomarkers, can increase the number of identified therapeutic options. We present a protein analysis of mutated sites and show that protein characteristics can provide potential indicators for the functional impairment of variants of unknown significance. Variant interpretation systems like Onkopus have the potential to significantly enhance the performance of personalized treatments, as they reduce the time required for variant interpretation and manual evaluation of personalized treatments, while maintaining reproducibility and traceability. We provide a free public instance of Onkopus at https://mtb.bioinf.med.uni-goettingen.de/onkopus.

## 1 Introduction

One of the current challenges for molecular tumor boards (MTBs) is the interpretation of genetic variants identified in the tumor of cancer patients. By assessing the pathogenicity and the clinical significance of these variants, molecular analysis provides new opportunities for the application of personalized therapies. Previous studies measured a mean time of 38.4 days before 2020 for molecular analysis and MTB discussion [1], indicating that variant interpretation is still mainly based on manual work and has great potential to be accelerated by automated interpretation, which can be confirmed by the authors’ clinical MTB practice. For the clinical interpretation of variants, various databases, such as Clinical Interpretation of Variants in Cancer (CIViC) [2] and OncoKB [3], have compiled data on the clinical significance of somatic variants. A challenge in the biological interpretation of variants is the pathogenicity classification, as they may cause severe damage to the protein or remain largely benign. While only a small fraction of variants has been classified as benign or pathogenic in a clinical study, the majority of variants remain variants of unknown significance (VUS). Machine-learning based methods to predict the pathogenicity of missense variants have been developed based on varying characteristics, including protein sequences and multiple sequence alignments [4, 5], conservation-based methods [6, 7], and methods based on varying different features [8–10].

An obstacle to the interpretation of variants in clinical practice is the necessity for manual effort to query numerous databases and aggregate the results. Variant annotation pipelines aim to close this gap by automating the process of annotating variant data. Multiple approaches have provided solutions for the biological annotation of variants [11–18]. Second, there are variant prioritization approaches focusing on the clinical interpretation of variants, providing classifications of the clinical significance of variants as well as therapeutic options [9,19–23]. To increase the availability and ease of use of variant interpretation, multiple approaches have presented interactive web front ends, which have focused on functional annotation [19, 24–27] or on cohort analysis [28]. However, the review carried out by Borchert et al. [29] on cancer variant interpretation tools revealed that no single tool was able to cover all knowledge bases required for all steps of variant interpretation equally.

Here we present Onkopus, an evidence-based variant interpretation framework supporting MTBs in the identification of pathogenic variants and the selection of personalized treatments for cancer patients. Onkopus provides both a customizable annotation pipeline for the functional annotation of variants, as well as the clinical interpretation of variants to support treatment decision-making in MTBs. It provides a web front end that enables users to query, upload, annotate and interactively explore genetic alterations individually and on the level of a patient molecular profile. We provide clinical annotations from different databases and show how aggregating searches for different match types can increase the number and suitability of therapeutic options for a molecular profile. We present methods for a detailed protein analysis of mutated sites, and show how protein characteristics, including surface accessibility, alpha carbon distances and protein domain affiliation, can serve as indicators for variant pathogenicity and a potential loss of function. Onkopus is available as a software package, an interactive web application, and workflows for the analytical framework KNIME [30].

## 2 Results

### 2.1 Overview of the Onkopus framework

Onkopus utilizes the advantages of a modular architecture: Each module is responsible for its own data and provides independent Application Programming Interfaces (APIs) endpoints, which follow a common pattern for syntax and semantic. This opens the option to query them individually or as part of the annotation pipeline (Fig. 1). Onkopus provides a comprehensive range of modules that encompass the full spectrum of variant characteristics, including transcript and protein information, population frequency, the clinical significance of variants regarding potential treatments, pathogenicity prediction, protein analysis and drug-gene interactions. Variant data is accepted in the form of VCF (Variant Call Format), MAF (Mutation Annotation Format) or CSV (Comma Separated Values) files or manual text-based queries for a direct variant search. A special feature of Onkopus is its ability to process the different variant nomenclatures from the biomedical context at DNA (e.g. ’chr12:g.25245350C*>*T’), transcript and protein level (’KRAS:p.G12D’) in HGVS [31] and VCF nomenclature. The process starts with annotating variants by identifying the variant type and mapping variant identifiers on transcript and protein level to DNA level (Extended Data Fig. 1). This is followed by the recognition of the mutation type, including single nucleotide variants (SNVs), copy number variations (CNVs), and gene fusions. Each variant type is associated with a list of module API endpoints (Table 1), which are consulted in order to interpret the variant. Onkopus classifies variants following the mapping of evidence codes to the Association for Molecular Pathology, American Society of Clinical Oncology and College of American Pathologists (AMP/ASCO/CAP) [32] guidelines proposed by MetaKB [33].

**Table 1:** Onkopus annotation scope: List of the supported mutation types and the associated annotation features in Onkopus.

| Type of the query | Category | Feature |
| --- | --- | --- |
| Single nucleotide variant | Genomics | HGNC gene symbol<br>Genomic location<br>CDS, UTRs<br>Liftover(GRCh37/GRCh38)<br>Known variants<br>Splicing variants |
|  | Population allele frequency | Total population frequency (dbSNP, gnomAD, ExAC) |
|  | Conservation scores | CADD, PrimateAI, GERP++, PhastCons |
|  | Protein sequence scores | AlphaMissense, ESM1b, Missense3D |
|  | Multi-feature scores (ML) | REVEL, MVP |
|  | Variant effect prediction | SIFT, PolyPhen2 |
|  | Clinical databases | ClinVar |
|  | Somatic databases | COSMIC |
|  | Transcripts | RefSeq, Ensembl |
| | Protein | AA exchange<br>Protein sequence<br>Protein structure prediction, PDB-IDs<br>Surface accession, $C_\alpha$ distances, secondary protein structure<br>Protein domains<br>Binding sites |
|  | Clinical evidence | Therapy evidence<br>Drug classification<br>Drug-gene interactions |
|  | AA features | Molecular weight, charge at pH7.4, aromaticity, polarity, phosphorylation, flexibility, solubility |
| CNA (In-Frame) | Pathogenicity | CADD, ESM1b |
|  | Transcripts | RefSeq, Ensembl |
| | Protein | Protein sequence<br>Surface accession, $C_\alpha$ distances, secondary protein structure |
|  | Clinical evidence | Therapy evidence |
|  | AA features | (see above) |
| CNA (Frameshift) | Clinical evidence | Therapy evidence |
| Fusion | Genes | Affected genes |
|  | Clinical evidence | Therapy evidence |
| Gene | Transcripts | RefSeq, Ensembl |
|  | Protein | Protein sequence |
|  | Clinical evidence | Therapy evidence |

**Figure 1:**
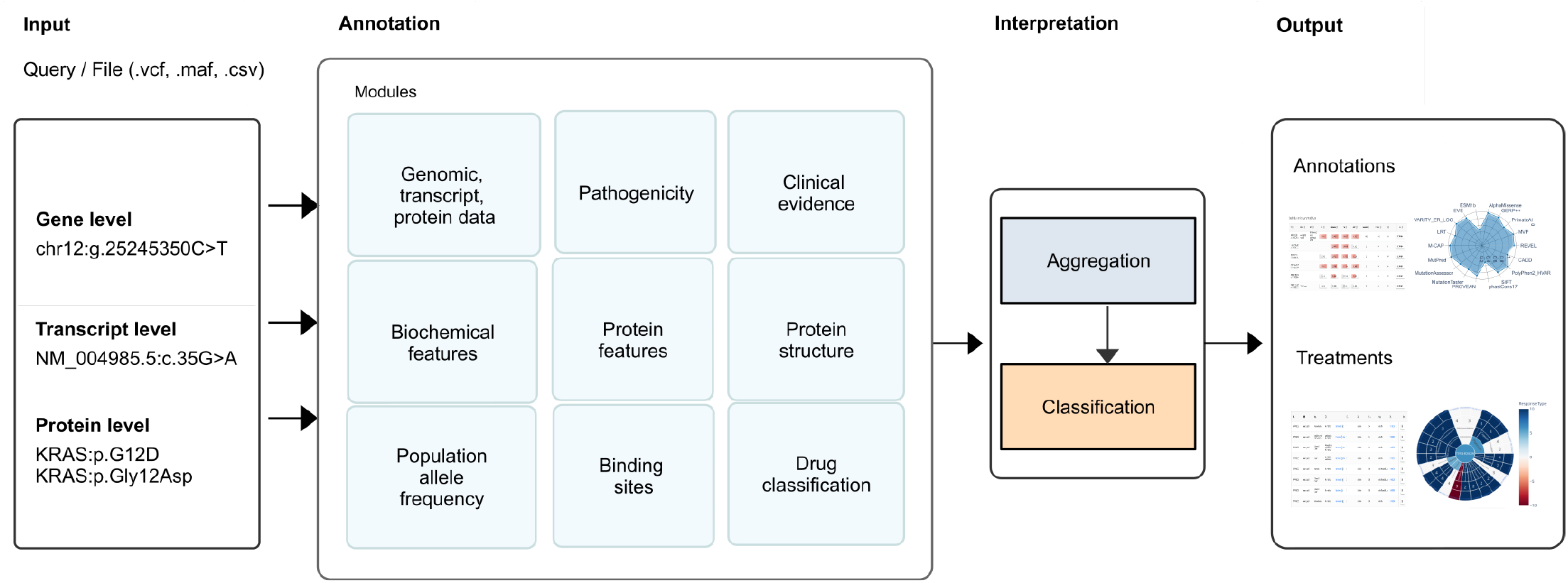
Onkopus architecture. The input can be a query or a provided variant file on DNA level, transcript or protein level. The annotation functionality is provided by the Onkopus modules, which may be consulted individually or as a full annotation pipeline. If the full annotation pipeline has been run, the retrieved data on the clinical significance of potential treatments is aggregated and classified. The output consists of lists of the annotated variants as well as potential treatments and their associated clinical significance. The results may be exported directly on the command line or interactively browsed in the web front end.

### 2.2 Functionality of the Onkopus web server

To make variant interpretation accessible to a broad community, we provide a publicly available software package and an interactive web front end. Variant data is provided by uploading a variant file in one of the supported formats, or by manually searching for biomarkers. Onkopus automatically detects the category of the provided variants, including SNVs, insertions and deletions, copy number alterations, gene fusions or genes, and lists the annotated variants in result tables listing selected features (Fig. 2a). For SNVs, we provide oncogene and tumor suppressor gene information, ClinVar reports, selected computational scores predicting variant pathogenicity, and the number of identified treatment options for a first impression of the clinical significance of the variant. This table is intended to provide an overview of a patient’s variants to identify promising variants for a further analysis on the linked detail pages. Besides of the variant table, we provide a clinical evidence table listing the clinical significance of the variants for potential treatments (Fig 2b). Both, the annotated variant data and their clinical significance regarding possible treatments are available as download as CSV files. Optionally, the user may choose which properties are to be exported (Fig. 2c). To facilitate the exploration of the variants, interactive plots are generated to illustrate the distribution of the variants on the chromosomes and the available treatment options (Fig. 2d).

**Figure 2:**
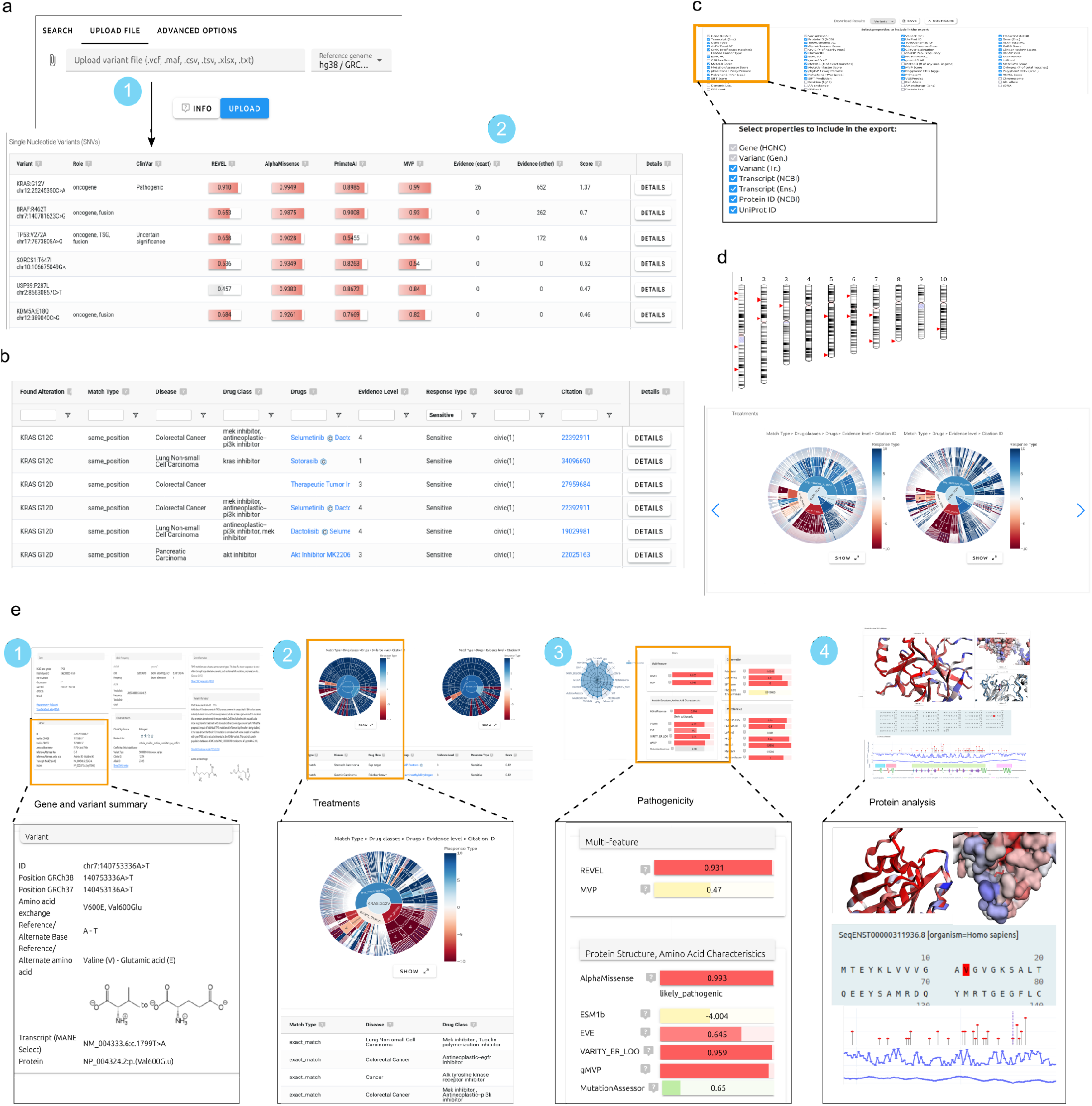
The Onkopus web front end. **a**, The input form allows to 1) enter a direct variant search or upload a file in one of the supported file formats (VCF, MAF, CSV). After the interpretation Onkopus presents 2) result tables listing the annotated variants for SNVs, InDels, gene fusions and genes ranked according to the clinical interpretation. **b**, A table retrieving potential treatments and the biomarker’s clinical significance, listing the associated biomarker, the match type, drugs, drug classification, evidence level, the response type and the citation ID of the underlying publication for each study. **c**, Customizable export features of the annotated variants and treatments, **d**, visualizations of the variant chromosomal locations and potential treatments, **e**, pages of the detailed view of a single nucleotide variant, including 1) genetic details, 2) clinical significance and potential treatments, 3) pathogenicity predictions and 4) protein analysis.

In addition, each variant can be analyzed individually on the linked detail page: For SNVs, we provide a comprehensive overview of details on genomic and transcriptomic characteristics, population allele frequency, clinical evidence, pathogenicity predictions and protein analysis (Fig. 2e). The tabs on the detail page reflect the steps of interpreting variants as it is practiced in MTBs like ours: 1) Obtaining general information about the gene and variant, 2) analyzing evidence-based treatments, 3) estimation of the variant pathogenicity, and 4) inspection of the mutation’s impact on the protein structure and characteristics. Similarly, we provide detailed pages for InDels, gene fusions (Extended Data Fig. 2c), and the entire gene. Each page provides persistent links that can be revisited at a later time or shared with colleagues for discussion of a variant.

### 2.3 A comprehensive overview of variant information

The first tab of the detail page provides some background about the selected variant, the corresponding gene and protein. We provide general information about the affected gene, including function, oncogene or tumor suppressor classification, the genomic location, transcripts, functional regions and the protein sequence. The general variant information provided includes data on population allele frequency, ClinVar reports, and CIViC summaries. In addition, we provide links to additional resources or information, including gene information (GeneCards [34]), variant information (CIViC), population allele frequency (dbSNP), and clinical reports (ClinVar).

### 2.4 Extended search increases findings of evidence-based treatments

To provide clinicians with an overview of potential treatments for a patient, we incorporated several databases collating clinical evidence data into Onkopus. We implemented separate modules for each database, thus enabling the possibility of future expansion to additional databases. One challenge in visually presenting treatment options for a patient is the necessity to consider multiple criteria, which clinicians evaluate when making a treatment decision. For instance, the classification of a biomarker may be of the utmost importance in an initial evaluation of potential therapy options. Nevertheless, further aspects may be of significant importance when looking at the clinical evidence in more detail, including the response type, the associated cancer type, or the associated biomarker. To facilitate this analysis of therapeutic options for a patient with the help of interactive visualization, we present, in addition to the evidence table, a sunburst visualization for the clinical interpretation of a variant on the detail page or a molecular profile on the overview page. The sunburst graph allows to display multidimensional data hierarchically spanning from the inner to the outer layers, where each layer represents a feature (Fig 3a). We constructed several sunburst graphs in order to facilitate the exploration of potential treatments according to different features, including visualizations based on drug classification, match type, cancer type and drugs.

**Figure 3:**
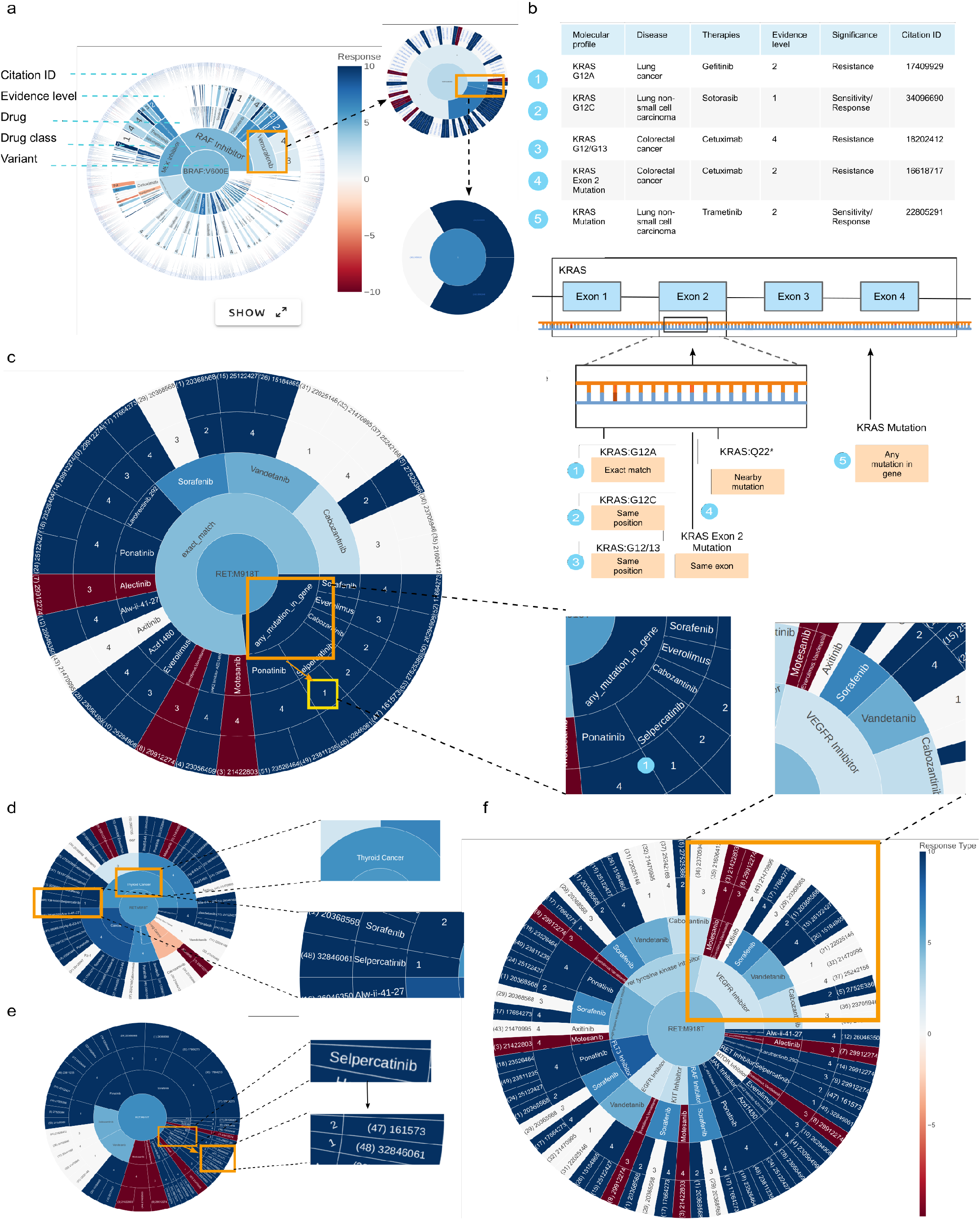
Clinical significance of variants on molecular treatments. **a**, Interactive sunburst visualization of the clinical significance of a biomarker regarding potential treatments. The sunburst visualization allows even large numbers of treatment options to be explored by clicking on each cell and triggering a zoom into the selected area. The outermost layer of each graph comprises the linked citation IDs, representing the number of evidences. The color coding represents the response type of the retrieved clinical evidence, with red indicating resistance, blue indicating sensitivity and white indicating unknown. **b**, Match types in Onkopus for the search for potential treatments for a specific biomarker, including 1) exact matches (e.g. ’KRAS G12A’), 2) evidence for variants with differing amino acid exchange at the same position (’KRAS G12C’), 3) variants at the same position with an arbitrary exchange (’KRAS G12’), 4) variants within the same exon or 5) clinical evidence on targeting the whole gene (’KRAS mutation’). **c**, Sunburst visualization of the retrieved clinical evidence for RET:p.M918T visualized according to the sequence ’match type’ *>* ’drug’ *>*, ’evidence level’ and ’citation ID’. **d**, RET:p.M918T sunburst graph visualized according to cancer type, making it easy to spot clinical evidence of the same tumor entity. **e**, RET:p.M918T visualization according to therapies **f**, grouped according to drug classification, whereby the magnitude of the area within the drug classification layer indicates the amount of clinical studies found for a drug target.

A challenge in retrieving variant interpretations from clinical evidence knowledge databases is the harmonization of the varying associated biomarkers underpinning the clinical evidence: For example, studies may examine clinical evidence of biomarkers on the basis of individual variants (e.g. ’NRAS Q61L’, ’KRAS G11 A12insGA’), variants in an exon (e.g. ’BRAF exon 15’), or biomarkers in the entire gene (e.g. ’NRAS mutation’, ’PCM1::JAK2’). As exon and whole gene associations may include point mutations and copy number alterations, we applied an approach that entails searching the integrated databases for varying biomarkers: In the first step, we search for exact matches in which the associated biomarker of the clinical evidence corresponds exactly to the variant of a patient (e.g. ’BRAF V600E’). In addition, we search for matches with differing base substitutions at the same genomic position (’BRAF V600R’), nearby mutations within the same gene (e.g. ’BRAF L597V’), mutations within the same exon (’BRAF Exon 5 mutation’), as well as any mutation in the gene (’BRAF mutation’) (Fig. 3b).

In order to aggregate the clinical evidence data from different databases, we defined a feature set with features available in all databases, including associated biomarker, cancer type, drugs, evidence level, response type, and the citation ID of the underlying study. Possible treatments for a patient are evaluated and presented based on these extracted features in Onkopus. All therapy suggestions in Onkopus are evidence-based and supported by at least one publication. We created a harmonization approach to unify the data contained in our included databases to directly compare treatment suggestions (Methods). For normalizing cancer types, we mapped the disease specifications of the source databases to the main tumor types defined by OncoTree [35] by combining exact and fuzzy matching (Methods). In order to assess the suitability of alternative treatments, it can be beneficial for the attending doctor to be aware of the primary mechanism of action of a given drug. We are thus retrieving drug classifications for the drugs of suggested treatments referring to the main target of a therapy.

We showcase the benefit of the Onkopus match types using the sunburst rays for finding clinical evidence on RET:p.M918T, a pathogenic variant that may occur primarily in thyroid and lung cancer tissues. For this specific variant, Onkopus yields several studies via exact match search, but only one study for Vandetanib reported with an evidence level of 1, and in this case the response type is not detected. In addition to the search for exact matches, the search for general mutations in RET as a biomarker identifies a study investigating Selpercatinib for patients with thyroid cancer, which was classified as level 1 indicating a sensitive response (Fig. 3c). The clinical evidence results may as well be explored according to the cancer type, allowing to specifically search for clinical evidence of the same tumor entity (Fig. 3d) or drug (Fig. 3e). In addition, the visualization based on drug classification provides an overview of the main target of each therapy, enabling MTBs to analyze the number of evidences for a drug target and to search for other therapy options (Fig. 3f). In case of RET:p.M918T, we can find multiple evidences for VEGFR inhibitors and RET tyrosine kinase inhibitors. In addition to the exploration of therapy options for a single variant, we can visually explore potential therapies for a patient by considering all biomarkers (Extended Data Fig. 3a) and summarized for the entire molecular profile (Extended Data Fig. 3b).

### 2.5 Aggregation of computational methods improves variant pathogenicity prediction

The impact of a variant on the protein is highly variable and can be estimated from a number of factors, including the specific amino acid substitution, the biochemical characteristics, the evolutionary conservation, or the mutated site within the protein. To predict the effect of sequence variants at the functional level, we included numerous computational methods for variant pathogenicity prediction. However, as these methods are based on different methods, their predictions may differ considerably. For this purpose, we provide a comprehensive variant annotation with numerous computational methods, including methods based on protein sequences including AlphaMissense [4], ESM1b [36] and EVE [37], conservation-based scores including PrimateAI [6], CADD [38] and GERP++ [7], functional effect prediction including SIFT [39] and PolyPhen2 [40], as well as machine-learning based methods based on different input features, including REVEL [8] and MVP [41], as well as other scores [7, 42–48]. In the web front end, we visualize the predictions yielded by the computational methods as score bars (Fig 2e(3)) showing the raw predictions score relative to the maximum values, and radar charts for direct comparisons (Fig. 4a). Information texts on each method are accessible via the information icons, ensuring that novice users have convenient access to guidance.

**Figure 4:**
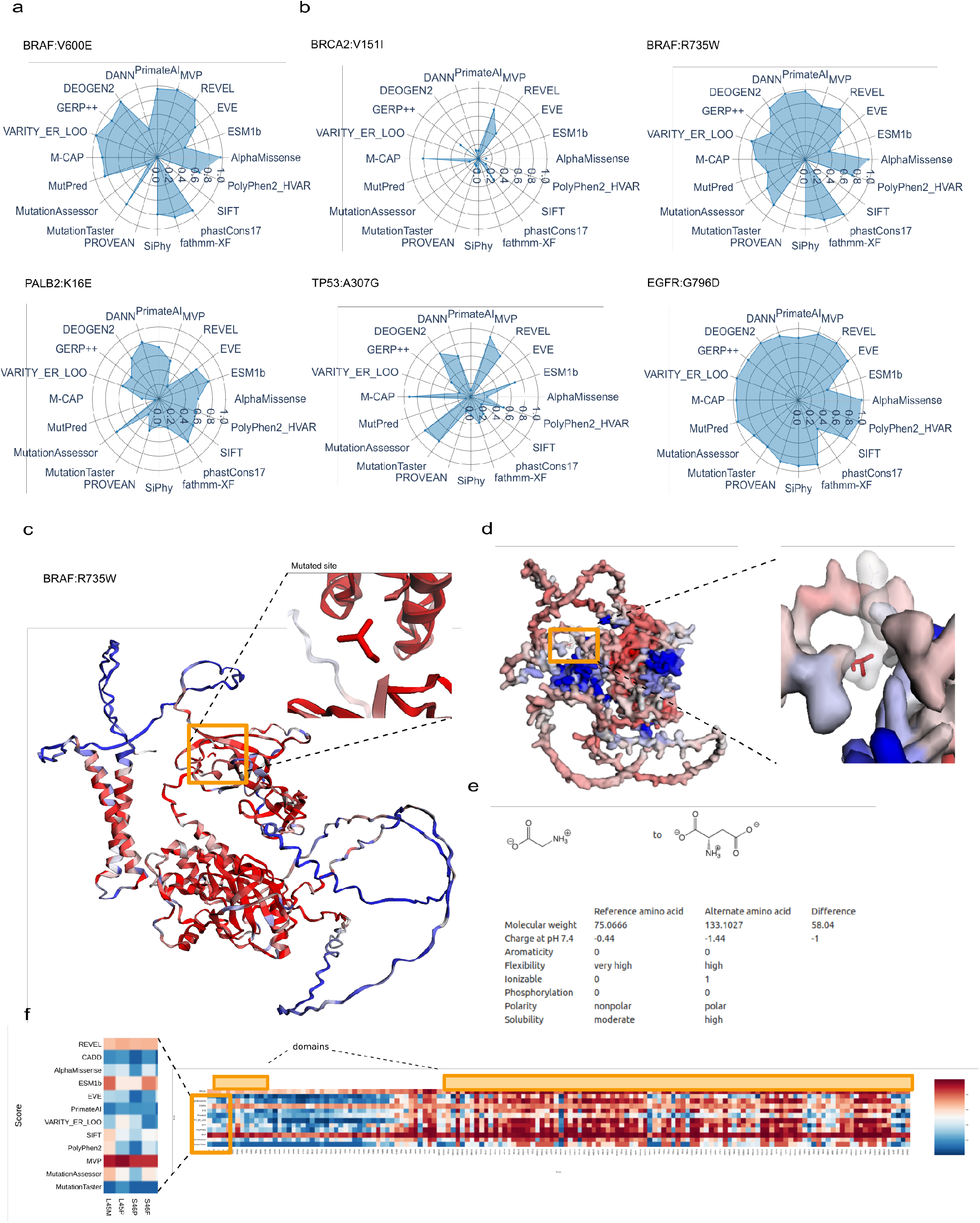
Variant effect and pathogenicity predictions. **a**, Comparison of pathogenicity predictions of computational methods included in Onkopus on selected missense variants reported as ’Pathogenic’ (BRAF:V600E) and ’Benign’ (PALB:K16E) in ClinVar, ranging from 0 (benign) to 1 (pathogenic). **b**, Selected variants reported as ’Unknown significance’ in ClinVar. In these examples, the majority of methods tend to predict BRCA2:p.V151I and TP53:p.A307G as rather benign, while BRAF:p.R375W and EGFR:p.G796D are predicted as significantly more pathogenic. **c**, Pathogenicity visualization in Onkopus at the protein level, colored by the average AlphaMissense-predicted pathogenicity at each site, with blue indicating benign and red indicating pathogenic. **d**, ScanNet-predicted binding sites, where the coloring matches the predicted binding site probability, with blue indicating low and red indicating a high probability. **e**, Comparison of the molecular amino acid features of glycine and aspartic acid, the wild-type and alternate amino acids of EGFR:p.G796D. **f**, Comparison of pathogenicity predictions of various scores included in Onkopus of TP53 tumor variants, ranging from benign (blue) to pathogenic (red).

When comparing these methods to predict the pathogenicity of variants, we can observe a consensus on the majority of scores in many cases for benign, pathogenic as well as loss-of function variants (Fig. Extended Data Fig. 4a, Extended Data Fig. 4b, Extended Data Fig. 4c). When comparing these methods to predict the pathogenicity of VUS, we can thus typically gain an overview of whether the variant is rather benign or pathogenic (Fig. 4b). In order to support the exploration of the mutated site within the context of the region of the mutated site, we have expanded the variant annotation with Onkopus to include protein information. We integrated interactive 3D visualizations of the predicted protein structure, indicating the pathogenicity (Fig. 4c) and predicted binding sites at each position (Fig. 4d).

### 2.6 Protein feature analysis support the interpretation of variants of unknown significance

The characteristics of amino acids may have a significant impact on the protein. For instance, the presence of an alpha helix breaker, including glycine and proline, may affect the protein structure and stability. To analyze the variant amino acid exchange at the molecular level of the protein, we provide an annotation with structural features of the reference and alternate amino acids, including differences in the molecular weight, charge, polarity, aromaticity, flexibility, ionization, phosphorylation and solubility (Fig. 4e). We illustrate the modification of the protein at the molecular level by comparing the biochemical properties of the wild-type and the substituted amino acid, as well as BLOcks SUbstitution Matrices (BLOSUM62) [49] scores of the amino acid exchange. In the case of TP53 tumor variants, we can identify contiguous areas that are classified as rather benign or pathogenic by the majority of scores (Fig. 4f, Extended Data Fig. 4d).

To further refine the analysis of pathogenicity at the protein level, we calculate key properties of the protein and the mutated site for variants. We calculated the relative accessible surface area (RSA) of each position within the protein, domain affiliations and the secondary protein structure. The distances of alpha carbon atoms within the protein were retrieved by calculating the distance of each amino acid’s alpha carbon atom relative to the mutated amino acid’s alpha carbon atom in 3-dimensional space (Fig. 5a). In the front end, we provide an interactive, aligned visualization of the protein, including the position of the mutated site and the distribution of nearby variants reported in the CIViC database on the sequence in the first row. The next rows visualize the surface accessibility, *C*_*α*_ distances, the protein domains, and the secondary protein structure (Fig. 5b).

**Figure 5:**
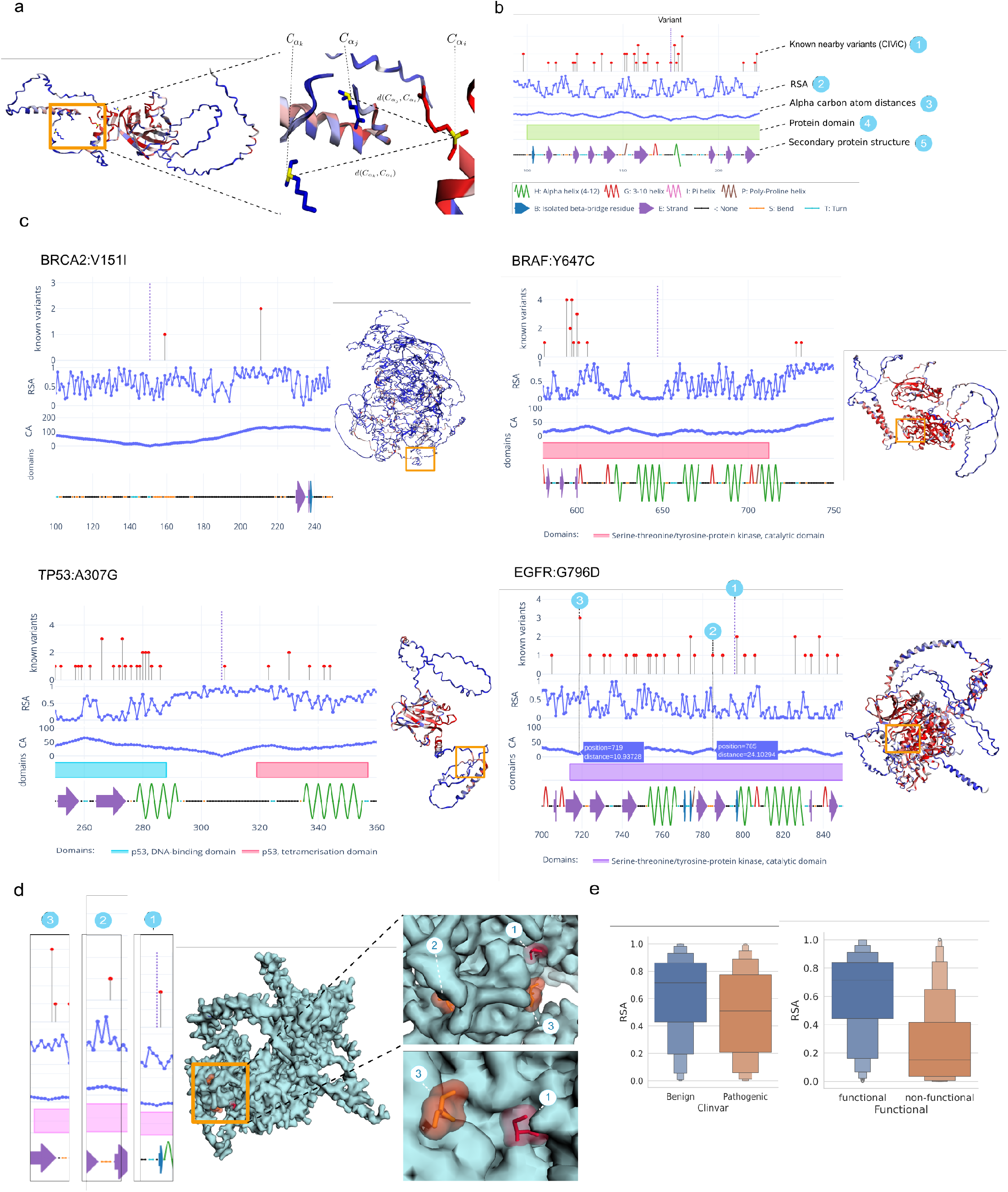
Protein feature analysis. **a**, *C*_*α*_ atom distances were retrieved by calculating the Euclidean distance of each amino acid’s alpha carbon atom relative to the mutated site’s alpha carbon atom, **b** Protein feature graph visualized in Onkopus web interface, including 1) number of known biomarkers included in CIViC at each position (mutated site marked as dashed line), 2) the relative accessible surface area, 3) the *C*_*α*_ distance of each amino acid to the mutated position, 4) protein domains and 5) the secondary protein structure. **c**, Protein features for selected variants of unknown significance (VUS): BRCA2:V151I and TP53:A307G are both located outside of protein domains and secondary structure elements and predicted as rather benign by AlphaMissense. BRAF:Y647C and EGFR:G796D are both located in protein domains and secondary stuctural elements, with low surface accessibility and a moderate to high AlphaMissense predictions, which may indicate pathogenicity. **d**, Structural view of EGFR:G796D and 2 biomarker positions within the protein with known clinical significance in CIViC, EGFR:T785A and EGFR:G719A/D/S. Although position 785 appears to closer to the mutated site due to its location within the amino acid sequence, the *C*_*α*_ distance calculation reveals that position 719 is in closer proximity to the mutated site 796 in the 3D structure. **e** The average RSA for variants from Clinvar (left) and the TP53 database. The values for benign or functional variants (blue) are compared with the pathogenic or non-functional ones (ochre).

For the analysis of a variant within the context of the protein, it is necessary to combine and interpret the protein features. While known variants from CIViC relate to a position on the sequence, the distances of the *C*_*α*_ atoms are more indicative of the 3D spatial relationship within the folded protein. Combined with the 3D protein viewer, it is possible to ascertain whether the VUS of the patient and a known variant, for example, are forming the same pocket. This can be confirmed through the annotation of the protein domains. The packaging of proteins is typically very compact, leading to a higher impact of the amino acid change in the center of the protein, which is represented by the RSA. Combined with the information of the amino acid exchange, e.g. size or polarity of the exchanged amino acid (Fig. 4e), the impact on the protein structure can be assessed.

To showcase how the protein analysis can help to interpret variants of unknown significance, we used the Onkopus protein analysis to examine selected variants that were reported as VUS in ClinVar (Fig 5c). As for BRCA2:V151I and TP53:A307G, the mutated positions are located in a loop structure and not affiliated with a protein domain or secondary protein structure element, while their RSA is higher than the average RSA of all amino acids within their protein. Both variants are predicted as rather benign by AlphaMissense. BRAF:Y647C is located within a protein domain, showing a low surface accessibility and a medium to high pathogenicity prediction score by AlphaMissense, indicating a rather pathogenic variant. EGFR:G796D is located within the catalytic domain within an Alpha helix structure, whereby the presence of multiple pathogenic variants in close proximity may be indicative of pathogenicity. The alpha carbon atom distances may help in identifying spatially adjacent variants: In the case of EGFR:G796D, multiple known variants are present in the immediate vicinity, including EGFR:T785A. In the folded protein within three-dimensional space, however, positions may be closer to the mutated site whose sequence-based position is further away, e.g. EGFR:G719A (Fig. 5d).

In order to investigate the impact of structural protein features on variant pathogenicity, we extracted variants from ClinVar and the TP53 database and calculated the average surface accessibility (Methods). The average surface accessibility of pathogenic variants was observed to be lower than the surface accessibility of benign variants (Fig. 5e).

### 2.7 Custom reproducible variant interpretation workflows

The increasing amount of data available for variant annotation enables ever more precise interpretation of variants, but also increases complexity of annotation pipelines. In addition, not all annotations are required in each scenario: For example, while the identification of personalized therapies is a priority for precision medicine, a biochemical research project may be primarily interested in molecular properties. As the modular architecture of Onkopus allows the modules to be queried on an individual basis, we are able to create customized annotation and filtering pipelines that are precisely tailored to the needs of each usage scenario. To facilitate the generation of customized variant annotation pipelines, we implemented graphical, adaptable workflows for the analytical framework KNIME. The workflows comprise a series of genome annotation nodes, each of which represent a distinct annotation, filtering or data parsing process. The nodes may be added or combined dynamically, with each workflow commencing with a read node of the required format and concluding with one or more write nodes.

For example, we showcase a workflow of a biomedical project annotating population allele frequency, subsequently filtering out variants that are commonly found in the population before proceeding with further annotation (Fig. 6a). The same process is repeated with variants that are located in non-coding regions and are classified as ”benign” in ClinVar. In the following, the variants are annotated with various annotation modules, including pathogenicity predictions, protein features and clinical evidence data. For the case of a cohort-based approach within a biomedical context (Fig. 6b), variants at protein level are mapped to the genomic level and merged in one dataset. The data is then annotated with characteristics of the mutated sites and altered proteins, including data on the functional regions, transcripts and their protein sequences, surface accession, alpha carbon distances and secondary protein structures. The annotated variants and data on the clinical significance of variants regarding potential treatments are then be exported in one of the supported formats, including annotated VCF and column-wise export of annotated variant data (Fig 6c) and evidence-based data on potential therapies (Fig 6d).

**Figure 6:**
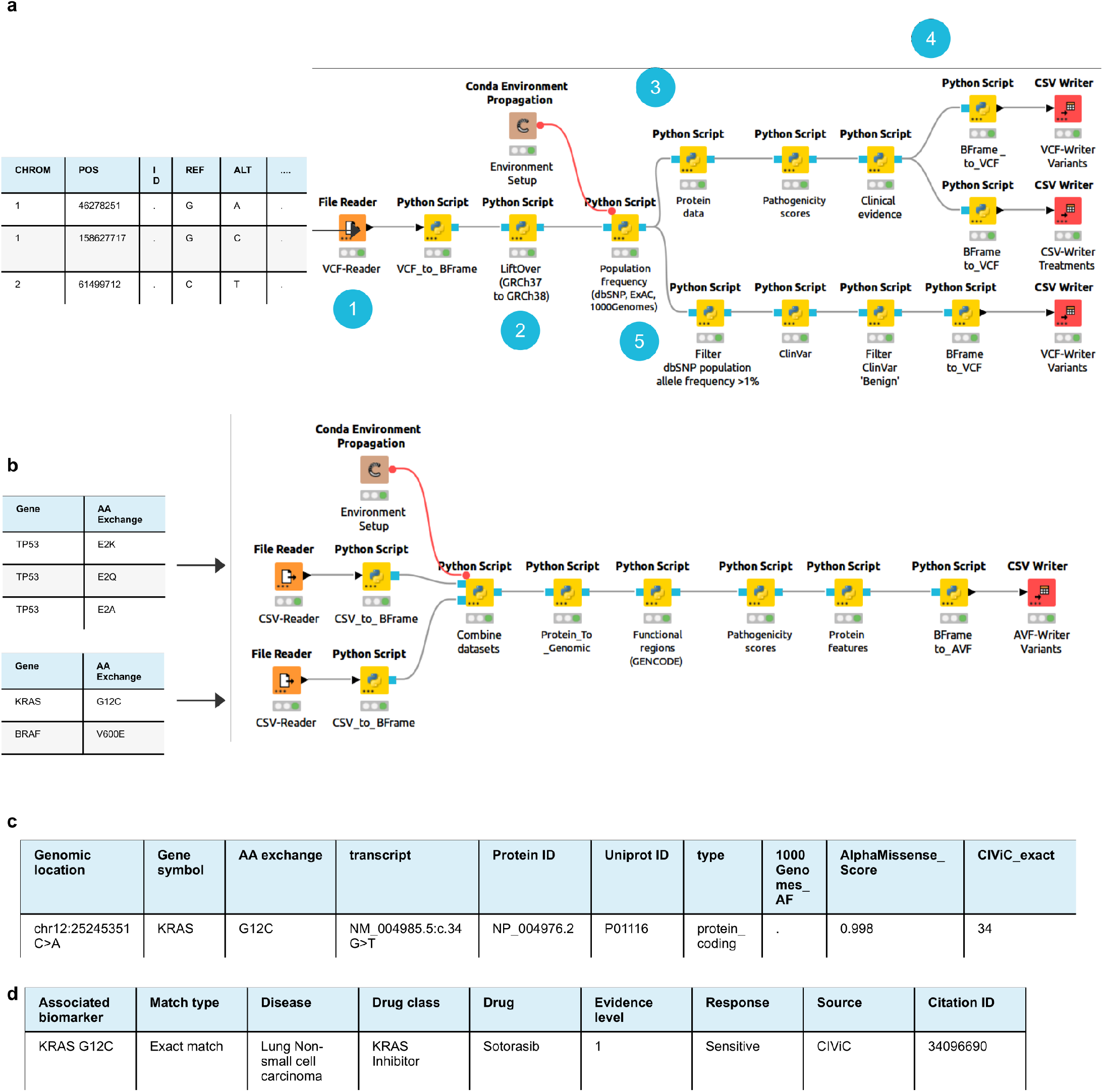
Custom variant interpretation pipelines using KNIME workflows. **a**, A variant interpretation pipeline to identify and annotate molecular cancer targets: 1) reading in variant data in VCF format, 2) converting it to GRCh38, 3) then annotating with population allele frequency. The annotation pipeline can be splitted up to generate different filtering pipelines as well, here the variant data is annotated and filtered according to variants that are common in the population and variant that are reported as ’Benign’ in ClinVar. Annotation modules are then added to the pipeline by sequentially arranging Onkopus modules, including pathogenicity and clinical evidence data of potential treatments. The annotated variants are then exported and annotated VCF format, the clinical significance of potential treatments is exported in CSV format. **b** A cohort-based pipeline retrieving biochemical data on proteins altered by SNVs: 1) Multiple datasets are read in and combined in one dataset. After annotating with functional regions, pathogenicity scores, and protein features, the annotated variants are exported in AVF and CSV format. **c** Example row of exported annotated variant data. Saved AVF files can be read in again, e.g. to annotate the variants later with additional modules. **d** Example row of the clinical significance data export

## 3 Discussion

Determining pathogenic variants in molecular profiles and selecting a targeted treatment is a main challenge in precision medicine. We propose Onkopus, a modular variant interpretation framework, as a powerful tool that supports MTBs in variant interpretation for selecting personalized therapies, as well as a dynamic software for annotating variants. Onkopus was built with the workflow of the MTB in mind: Special focus was set on providing comprehensive information to the one who is preparing the MTB in a clear manner. The user is not required to visit numerous different databases and websites but has all the information on hand and can focus on finding the best matching treatment for every patient. Following a search, the process starts with an overview about all searched variants, followed by details about evidence-based studies, and an analysis that supports the interpretation of VUS. As more and more methods and biomarkers will follow the route from translational research to standard therapies, the number of therapeutic options for personalized treatments will continue to grow. The modular architecture of Onkopus allows for adding new features to the Onkopus framework easily to follow the state of the art, with the web interface for the MTB as well as for the KNIME workflows for biomedical research. By the use of separated modules, it is possible to only install the main Onkopus package and access the publicly available Onkopus modules without the need to download large software packages. On the other hand, it is possible to download the entire framework and databases and set it up in environments with limited or no internet access, such as in a clinical environment. Targeted annotation with selected modules can therefore help researchers to annotate variant data faster and in a more targeted way, while only the modules that are really needed need to be installed locally. In addition, the distributed architecture makes it easy to update modules or add new modules to the framework. Onkopus is able to parse variant data in numerous data formats and nomenclatures. This is particularly useful as variants are usually referred to at the protein level within MTB discussions (e.g. ’BRAF:V00E’), whereas the data in VCF files is at the DNA level. Clinicians thus have the option of interpreting a VCF file of a patient in a first step, as well as manually searching for a specific variant at a later time or during a MTB discussion.

An important aspect of the front end is the ability to view the clinical interpretation of a variant individually as well as in the context of the overall molecular profile. In this way, we provide MTBs an overview of all treatment options for a patient, while the sunburst visualisation makes it possible to search them interactively using various criteria. Since annotation requirements can vary greatly depending on the use case, we provide an easy-to-use solution within the KNIME workflows to create customized variant annotation and filtering pipelines. By searching for clinical significance based on the Onkopus match types, we are confident of increasing the number of potential treatments presented to clinicians while maintaining clinical significance can contribute to the selection of personalized treatments.

While computational methods show high accuracy in predicting the pathogenicity of missense variants, it has been pointed out that a single score cannot capture the complexity of the mechanisms of pathogenicity [41], and as the available methods are based on different data and have different foci, they have different strengths and weaknesses and may differ from one another [50]. A reliance on a single or limited number of methods for assessing pathogenicity within a MTB may result in a biased assessment of a variant. We thus consider it as an important step to provide a comprehensive overview of a variant, including different pathogenicity scores, as well as the characteristics of the amino acid sequence and protein structure, towards understanding the biochemical processes involved in the pathogenicity of variants. A unique feature of Onkopus is thus its focus on providing the opportunity to analyze and interactively explore the entire protein of mutated sites. By combining the protein analysis with computational methods for predicting pathogenicity, MTBs can use pathogenicity scores to retrieve an initial assessment of the pathogenicity of a variant, followed by an extended analysis of the surrounding region of the mutated site. We are confident that the examination of a variant within the context of the entire protein is an effective method for elucidating the impact of the mutation and to ascertain the underlying mechanisms that contribute to its pathogenicity.

Although a large number of aspects of variant interpretation were considered in Onkopus, some limitations have to be pointed out: Onkopus is developed during a time where MTBs are still under development. The amount of data differs from organ tumor boards. While also more and more data get publicly available, annotated data in databases are not always structured, or need to be harmonized to bridge different semantics. The information stored in the included databases is not always consistent and presents a challenge to parse, particularly with regard to the response of patients to therapies. It is also important to consider that the calculated protein features are based on computationally predicted structures, which may contain errors. We do not evaluate the quality of the studies that support the evidence-based clinical interpretation. It is thus incumbent upon the clinicians to undertake a detailed examination of the proposed publication that supports the clinical significance of a biomarker. Furthermore, it is important to note that the harmonization of disease types based on a fuzzy matching approach does not guarantee the accuracy of all cancer type matches. Nevertheless, this approach has the advantage of reducing the extensive information on associated cancer types contained in clinical databases to a concise and comparable list of cancer types.

We are confident that the dynamic architecture of Onkopus will allow for a variety of dynamic usage scenarios for variant interpretation, including the support of MTBs and genome annotation for biomedical research. This potential offers the possibility of establishing Onkopus as a widely utilized solution for variant interpretation. Onkopus is designed as a long-term project as the MTB evolves and new biomarkers, methods and treatments are being accessible to the clinic. Additional modules will be added in the future and existing modules are regularly updated.

## 4 Methods

### Collecting genomic, transcript and protein information

Onkopus retrieves data on the genomic, transcript and protein characteristics of genetic alterations by employing SeqCAT [51]. For variants on genomic level, SeqCAT is queried to retrieve the HGNC gene symbol, the amino acid exchange, the transcript ID and the protein ID of the RefSeq MANE Select transcript. Variant data on protein level is mapped to genomic level by matching the gene symbol and amino acid exchange into gene coordinates based on the RefSeq MANE Select transcript. Functional regions, including gene start and end positions, all transcripts of a gene, exons, coding sequences and untranslated regions (UTRs), the GENCODE human release v44.2 [52] annotation was downloaded from https://ftp.ebi.ac.uk/pub/databases/gencode/Gencode_human/. Data on cancer driver genes has been extracted from the COSMIC v99 Cancer Gene Census (CGC) [53], which was downloaded from https://cancer.sanger.ac.uk/census.

### Collecting data on population allele frequency

For retrieving data on allele population frequency, the dbSNP frequency database [54] was downloaded from https://ftp-trace.ncbi.nih.gov/snp/population_frequency/latest_release/. For retrieving data from gnomAD [55] and 1000Genomes [56], dbNSFP [57] v4.5a was retrieved from https://dbnsfp.s3.amazonaws.com/dbNSFP4.5a.zip.

### Collecting data on variant pathogenicity

For clinical variant assessments, the ClinVar [58] database was downloaded from https://ftp.ncbi.nlm.nih.gov/pub/clinvar/. To collect data on the pathogenicity of variants, multiple modules were created for methods on variant pathogenicity prediction. AlphaMissense scores were downloaded from https://zenodo.org/record/8208688, REVEL scores were downloaded from https://rothsj06.dmz.hpc.mssm.edu/revel-v1.3_all_chromosomes.zip, LoFTool scores were downloaded from https://academic.oup.com/bioinformatics/article/33/4/471/2525582. Additional scores were extracted from the dbNSFP database. For score values that contain several values separated by semicolons in dbNSFP, we select the maximum value in each case. For direct score comparisons within the pathogenicity radar visualizations, dbNSFP rankscores were extracted.

### Computing protein structures and features

To compute different features of a mutated site’s protein, we predicted wild-type protein structures using OmegaFold [59]. The relative accessible surface area (RSA) and the secondary protein structure were computed with DSSP (Dictionary of Protein Secondary Structure) [60, 61]. The RSA [62] of a protein residue quantifies the proportion of the protein’s surface that is exposed to the surrounding environment (Accessible surface area (ASA)) compared to a fully extended reference state (Maximum possible solvent accessible surface area (MaxASA), calculated as

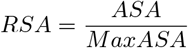

The *C*_*α*_ distances were calculated using the Euclidean distance between each amino acid’s 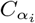 atom and the 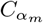 atom position of the mutated amino acid:

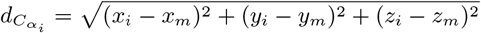

Protein domain information included in the Pfam database [63] has been retrieved using InterproScan 5 [64]. BLOcks SUbstitution Matrices (BLOSUM62) scores representing alignments between evolutionary divergent protein sequences were computed using the Python blosum package. PDB (Protein Data Bank) files were parsed using the BioPandas python package [65]. For computing the biochemical characteristics of amino acids, Biopython [66] was employed. Protein binding sites were predicted using ScanNet [67].

### Collecting clinical variant interpretation knowledge

To retrieve data on the clinical significance of biomarkers regarding possible treatments, we implemented modules for the CIViC [2], OncoKB [3] and MetaKB [33] databases. The CIViC database files available at https://civicdb.org/releases/main were downloaded. For OncoKB, the public Application Programming Interface (API) endpoint available at https://www.oncokb.org/api/v1/annotate/mutations/byGenomicChange is queried. For authorization, the OncoKB module requires an authorized OncoKB API key included in the HTTP header of an active OncoKB account in order to query the public API. For MetaKB, publicly available JSON files were downloaded from https://drive.google.com/drive/folders/1ZY6o3uaLOZSjOQPpXSMsnMbXmFWpb58d). To enable fast requests, the files are loaded as dictionaries, where for each entry we extracted the associated biomarker, gene, variant exchange, response type, evidence statement, disease, and citation address. To enable queries for exact matches of the MetaKB databases, dictionaries were created based on the associated biomarkers. Requests for the whole gene were enabled by creating a dictionary based on the associated gene. To aggregate the clinical evidence entries of the integrated databases, we extracted the following set of features extracted for each entry: Associated biomarker, cancer type, drugs, response type, evidence level, evidence statement and citation ID.

To normalize evidence levels between databases, the harmonization approach proposed by [33] for evidence level harmonization was employed. To harmonize the proposed treatments, CIViC treatments with multiple drugs whose therapy interactions were labeled as ’substitutes’ were converted into separate treatment options. To normalize cancer types, we extracted the main tumor types defined by OncoTree [35]. The disease specification of the entries from CIViC, OncoKB and MetaKB were converted to lowercase and common synonyms were normalized. We then applied a combination of exact and fuzzy matching to the OncoTree cancer types. The method first checks for an exact match between the source data and the standardized cancer types. If no exact match is found, the method applies fuzzy matching using the token sort ratio algorithm, which performs a fuzzy comparison to measure similarity, and selects the highest scored cancer type term. The token sort ratio *R* is computed as the Levenshtein distance *D*(*S*1, *S*2) between the token strings *S*1 and *S*2

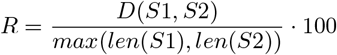

To normalize the data regarding the sensitivity of targets, we generated a custom mapping for each database. For CIViC, all values from the ’significance’ column were extracted that contain ’Sensitivity’ or ’Resistance’. For OncoKB, the response types ’R1’ and ’R2’ were mapped as resistant, and ’1’, ’2’, ’3A’ and ’3B’ as sensitive. As for MetaKB, the descriptive value of the evidence association was mapped as response type. To merge duplicates among clinical evidence databases, treatments were merged if the normalized features were identical (drug label, cancer entity, evidence level, response type, citation ID).

### Computing drug classifications

Drug classifications were retrieved from DrugOn, a drug classification system based on manual and automated classifications [68]. The automatic process accesses information from public resources, including DrugBank [69], ATC codes and MeSH terms.

### Building modules and API endpoints

The majority of the Onkopus web services were built using Python and the Flask web framework. Table-formatted databases, including dbNSFP, REVEL and CIViC, were indexed using tabix [70]. For database files based on CSV and GTF format (GENCODE, COSMIC, DGIdb), SQL-based database tables were generated from the data files using the ’LOAD DATA IN FILE’ SQL command. In order to accelerate data queries, indexes were then created on the relevant columns of the database tables. All modules have a preprocessing script that automatically downloads the requisite databases and generates the necessary data at startup. This ensures that the installation and use of Onkopus is also possible in environments without limited or no internet access, such as in a clinical infrastructure. The annotation procedure has been accelerated by parallelizing module requests. In order to permanently store molecular profiles of patients, we have created a database module based on MariaDB that stores the identified biomarkers for each patient. Dockerfiles [71] are provided for each module, all modules can thus be started locally or as Docker containers. OpenAPI (swagger) documentation is provided for all API endpoints to support the development of specialized client applications.

### Data parsing and variant normalization

Variant data parsing, reading and writing files, variant validation and normalization in HGVS and VCF notation, variant type recognition, and liftover transformations between genome assemblies were realized using AdaGenes. Onkopus works internally on hg38 (GRCh38), other reference genomes, including hg19 (GRCh37) and T2T-CHM13, are utilized by using LiftOver [72]. The AdaGenes biomarker frame was employed as the base data structure for all Onkopus modules, serving as both input and output data format for each module client. Variant data on transcript or protein level is mapped to DNA level by retrieving the genomic positions using SeqCAT. To parse variants in text-based user queries, AdaGenes was used to identify biomarkers in different nomenclatures. The annotation of variant data at transcriptome or protein level is made possible by mapping the data to genome level by employing SeqCAT.

### Module clients

As the number of characters transmitted via HTTP GET requests is limited, the Onkopus clients split the variant list into subsets and send separate requests for each variant list, finally the results are aggregated again.

### Web front end

The web application (Onkopus Web) was built using the Vue.js Javascript framework, layout formatting has been implemented with the Vuetify package. Interactive tables were implemented using AG Grid. Interactive radar plots, sunburst plots and protein feature visualizations were implemented using Plotly [73]. Chromosome locations were plotted with Ideogram.js. 3D protein structures were visualized using 3DMolJS [74]. Genome sequencing data was visualized by integrating the IGV genome browser [75]. Vector images of the protein and sunburst graphs were generated using Python Kaleido. Requests from the front end are processed by an additional module, the Onkopus Server. The server module provides API endpoints aggregating all Onkopus modules into a unified response and is employed when a complete annotation with all modules is performed. The server initiates requests to the modules and subsequently aggregates the responses into a unified output. In addition, the server module provides API endpoints for the interactive visualizations. The API documentation is available at https://mtb.bioinf.med.uni-goettingen.de/onkopus/api/v1/.

### Analytical Workflows

To generate customizable graphical variant annotation pipelines, we prepared workflows for the Konstanz Information Miner (KNIME) [30]. Each KNIME node is coupled to an Onkopus module and consults its API endpoints to retrieve annotation data. The input and output of each node is a nested JSON data structure, which is annotated or transformed by each node depending on its functionality. For parsing variant data from VCF, MAF or CSV files, as well as to save variant interpretation results to files, nodes from the AdaGenes package have been integrated.

### Benchmarks on public datasets

To test the parsing and processing of genetic mutation data, TCGA MAF files were downloaded using the TC-GAbiolinks package [76]. For analyzing features of known benign and pathogenic variants, we extracted variants from the ClinVar dataset (state: 2024/03/07) that were labeled as ’Benign’ or ’Pathogenic’ and were identified as protein-coding variants by SeqCAT, resulting in a dataset of 1904 variants labeled as benign, and 4403 as pathogenic. Tumor variants of TP53 [77] were downloaded from https://storage.googleapis.com/tp53-static-files/data/ TumorVariantDownload_r20.csv https://tp53.isb-cgc.org/get_tp53data. We extracted all variants that were labeled as ’functional’ or ’non-functional’, resulting in a dataset of 508 variants labeled as functional and 445 variants labeled as non-functional.

### Code availability

The source code of all Onkopus modules is available at https://gitlab.gwdg.de/MedBioinf/mtb/onkopus, the main Onkopus Python package is available at https://gitlab.gwdg.de/MedBioinf/mtb/onkopus/onkopus. Onkopus is available as a Python module via PyPI (https://pypi.org/project/onkopus) or Anaconda (https://anaconda.org/anaconda/onkopus). Pre-built Docker images of all Onkopus modules are available on Docker Hub (https://hub.docker.com/nadinekurz/onkopus). A public instance of Onkopus Web is available at https://mtb.bioinf.med.uni-goettingen.de/onkopus. The Onkopus KNIME workflows are available at the KNIME Hub https://hub.knime.com.

## Data Availability

All data produced in the present study are available https://gitlab.gwdg.de/MedBioinf/mtb/onkopus.

https://portal.gdc.cancer.gov/

https://civicdb.org/

https://www.oncokb.org/

https://search.cancervariants.org/

https://www.ncbi.nlm.nih.gov/clinvar/

https://sites.google.com/site/jpopgen/dbNSFP

https://www.ncbi.nlm.nih.gov/snp/

https://www.gencodegenes.org/

https://cancer.sanger.ac.uk/cosmic

## Acknowledgements

We thank the molecular tumor board of the University Medical Center Göttingen, including Li Beißbarth, Raphael Koch, Tobias Overbeck, Julia Beck, Nelia Nause and Kirsten Reuter-Jessen, for providing our team access to the MTB conferences and providing support, substantive debates and feedback. We thank Charlotte Höltermann for designing the Onkopus logo. We also acknowledge the International Max Planck Research School for Genome Science (IMPRS-GS) for supporting NSK.

## Author contributions

Conception and initiation of the project was done by JD and TB. The architecture of Onkopus was developed by JD and NSK. The Onkopus main package was implemented by NSK. The Onkopus modules were implemented by NSK, KK, TT and KD. The web application was implemented by NSK and TT. The KNIME workflows were implemented by NSK and KK. Feedback and evaluation of medical use cases was contributed by AK. Testing environments for the public instance were set up by KD. The manuscript was written by NSK and JD, reviewed and approved by all authors.

---

**Extended Data Figure. 1:**
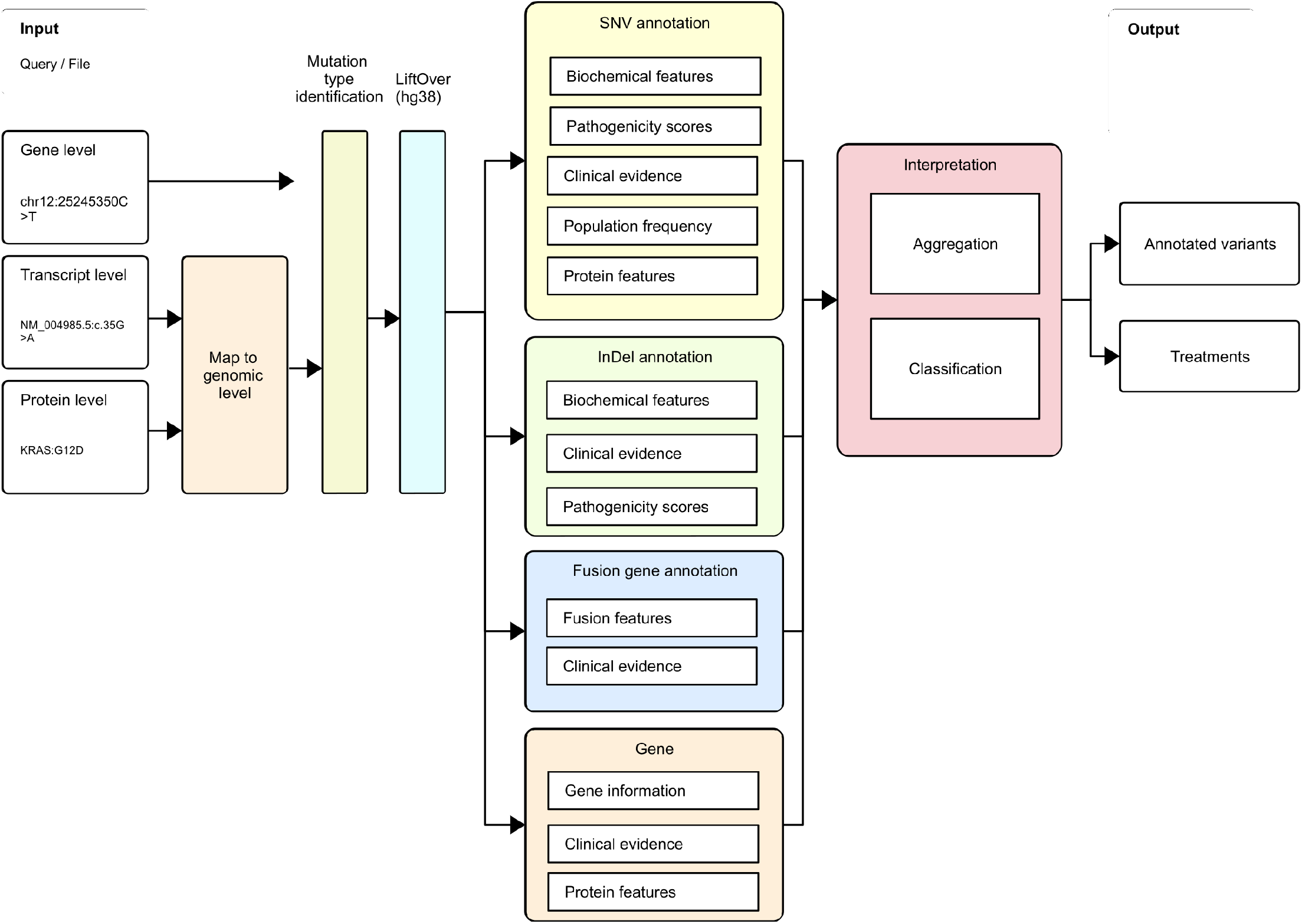
Onkopus annotation workflow. Complete annotation and interpretation pipeline for different DNA mutation types in Onkopus. Variant data on transcriptome and protein level is mapped to genomic level based on the MANE Select transcript. Variant data in hg19 is lifted over to hg38, then the mutation types of each variant is recognized. Based on the mutation type, Onkopus annotates the variants with the associated modules, then aggregates and scores the results.

**Extended Data Figure. 2:**
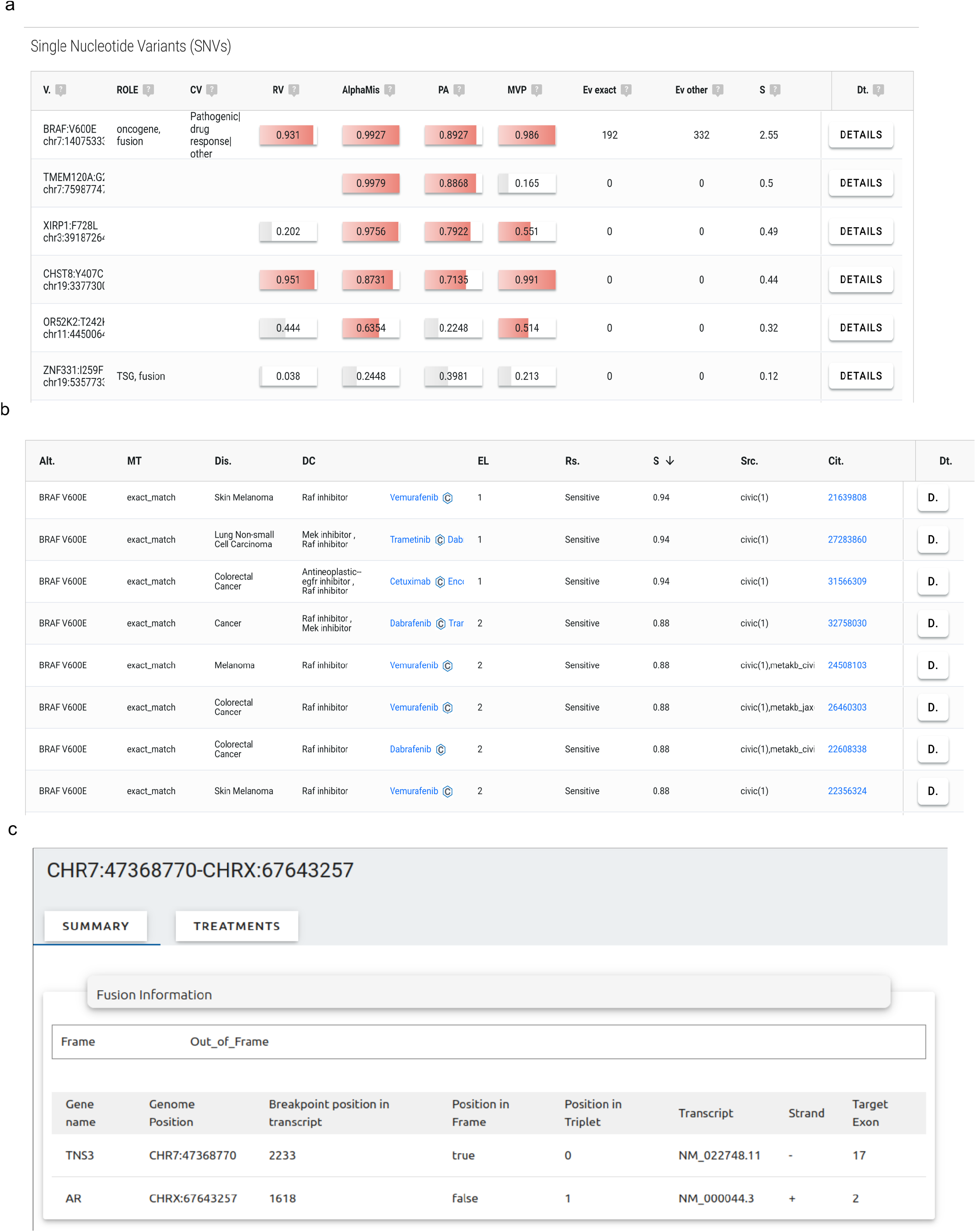
Variant interpretation in Onkopus Web. **a**, Onkopus presents the molecular profile as a list of variants, including the associated protein location, the COSMIC assessment, computational pathogenicity predictions, including REVEL, AlphaMissense, PrimateAI and MVP, as well as the number of retrieved exact match interpretations, and the overall number of interpretations. **b**, A list of the clinical evidence of possible treatment for a cancer patient, listing the associated biomarker, match type, cancer type, drug class, drug, evidence level, response type, the source database, as well as the citation ID of the underlying study, including a link to the publication.

**Extended Data Figure. 3:**
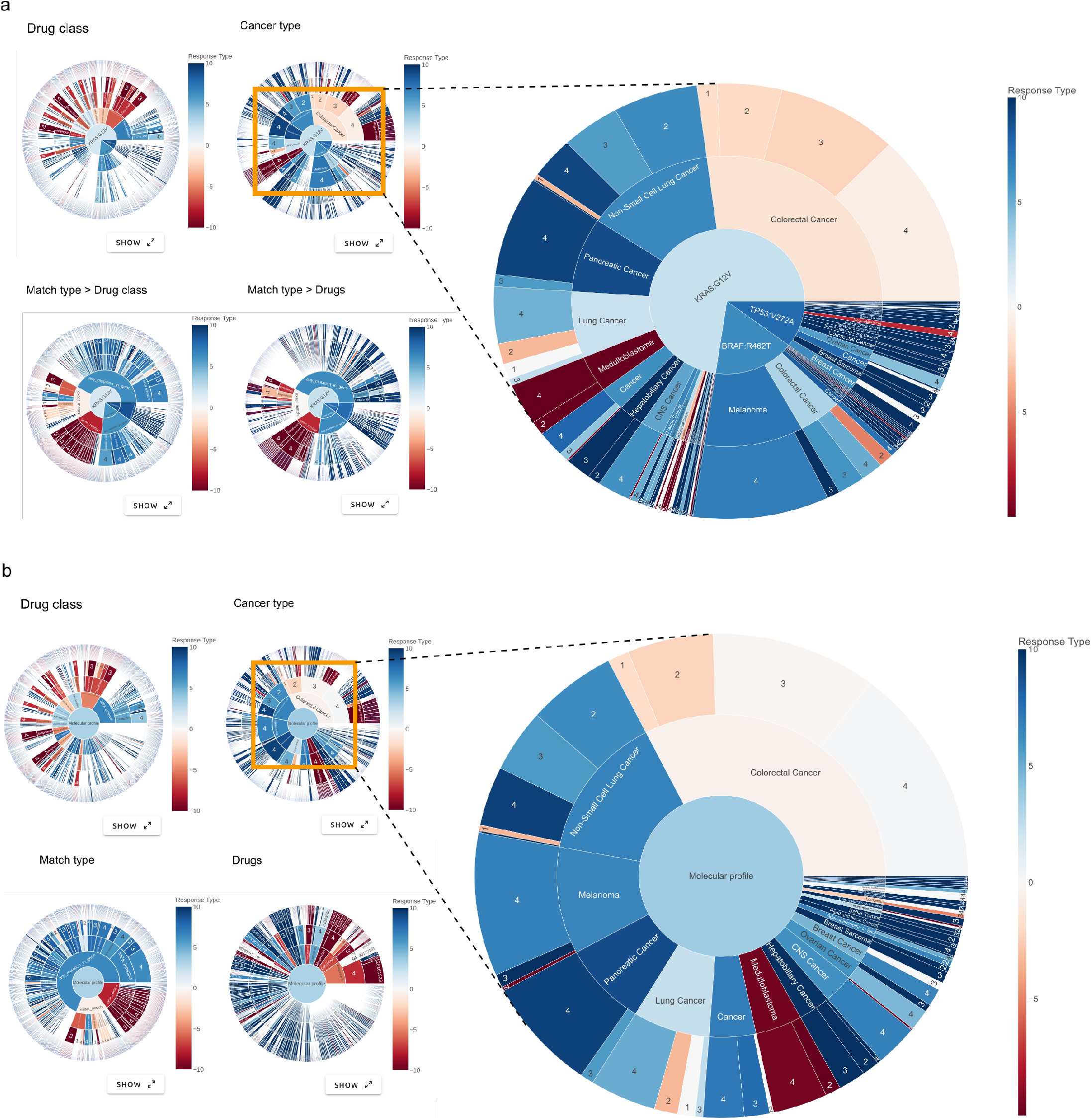
Visualization of treatment options in Onkopus on molecular profile level. **a** Sunburst visualizations of uploaded include clinical significance data of all variants of a patient, visualized according to drug classes, cancer type, drugs and match type. **b** Aggregated view of treatment options for a patient. As the number of clinical studies increases, so too does the size of the sunburst graph beams, which allows for straightforward analysis of the clinical evidence for each therapy, therapy class, cancer type and match type.

**Extended Data Figure. 4:**
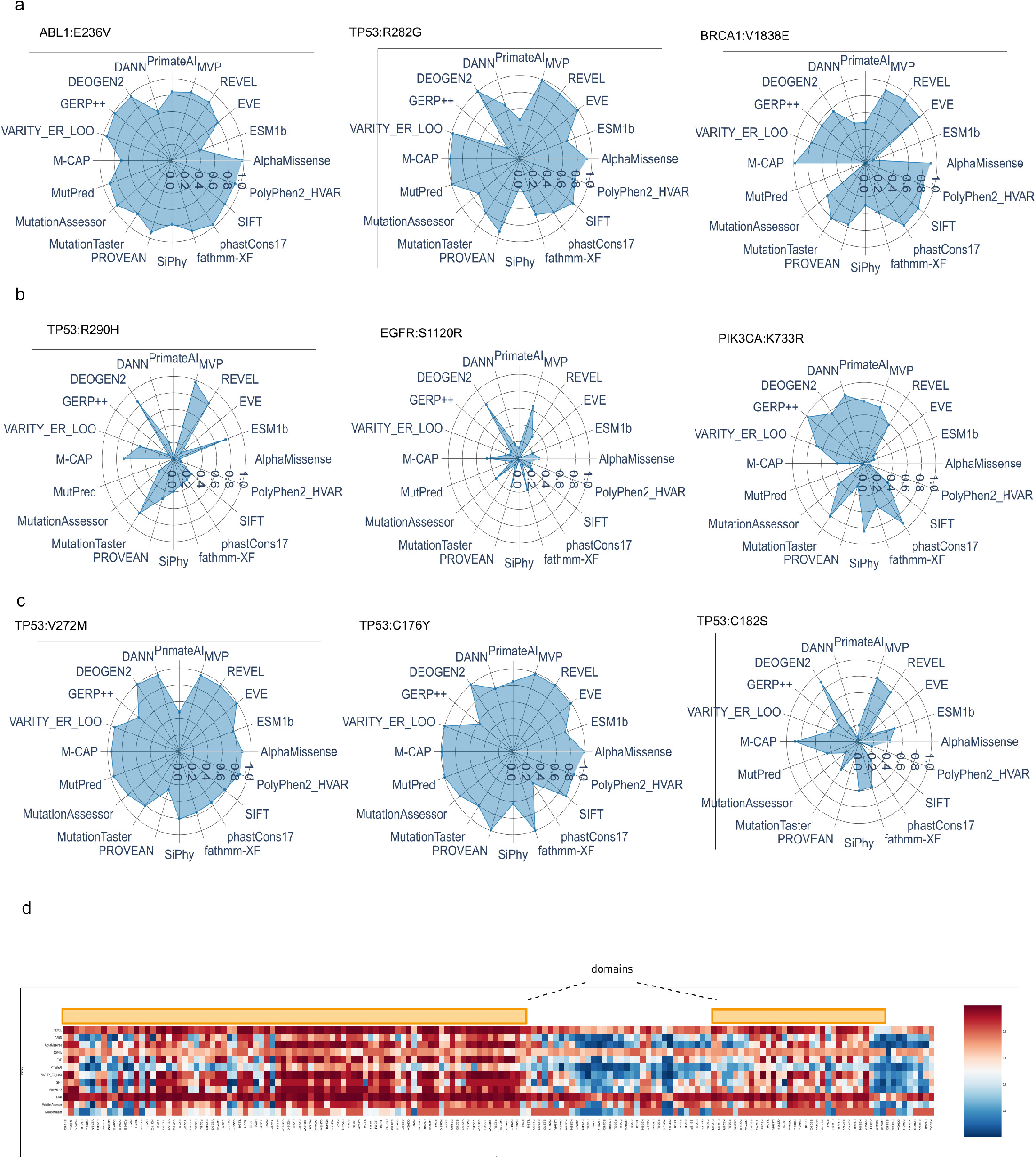
Comparison of pathogenicity predictions. **a**, Variants reported as pathogenic in ClinVar, **b**, variants reported as benign. **c**, Comparison of TP53 tumor variants reported as non-functional (TP53:p.V272M, TP53:p.C176Y) and functional (TP53:p.C182S).

